# *APOE* and *TREM2* variants differentially influence glial fibrillary acidic protein and neurofilament light in plasma of UK Biobank participants

**DOI:** 10.1101/2025.02.24.25322783

**Authors:** Yun Freudenberg-Hua, Luca Giliberto, Cristina D’Abramo, Wentian Li, Yilong Ma, Alison Goate, Jeremy Koppel

## Abstract

Plasma levels of protein biomarkers glial fibrillary acidic protein (GFAP) and neurofilament light (NEFL) are key dementia biomarkers, but it is unclear how risk genes for Alzheimer’s disease (AD) influence levels of these biomarkers. We investigated the association of the established high-effect variants for AD in *APOE* and *TREM2* with these biomarkers, using data from over 50,000 participants from the UK Biobank (UKB). The results show that *APOE4* is associated with elevated levels of plasma GFAP, and to a lesser extent, NEFL. The *APOE4* effect on GFAP increases with age and the number of *APOE4* alleles. The risk variants R47H and R62H in *TREM2* are associated with higher NEFL levels, but not with GFAP, and the effect sizes do not increase with age. Higher levels of both GFAP and NEFL in midlife are significantly associated with greater risk for incident dementia. In contrast, the protective *APOE2* allele showed no effect on GFAP or NEFL. In conclusion, we find that major genetic risk factors for AD differentially affect dementia protein biomarkers across age, indicating gene specific pathways with potential therapeutic implications.

## Introduction

Blood-based biomarkers for Alzheimer’s disease (AD) and related dementias have the potential to advance our understanding for brain pathology and present a minimally invasive and cost-effective approach to aide diagnoses, prognostication, and identification of at-risk individuals for early intervention(1). Two established blood protein biomarkers for dementia — neurofilament light (NEFL or NfL) and glial fibrillary acidic protein (GFAP)(2-4) — were measured in the UKB Pharma Proteomics Project, which aims to provide insights into proteomic signatures for health and disease(5). Both biomarkers were found to be highly predictive of incident all-cause dementia in UKB participants(6). NEFL is a biomarker of axonal damage and rising levels of plasma NEFL indicate an acceleration of neuronal injury and brain volume loss even in cognitively unimpaired older individuals(7). In contrast, GFAP is a biomarker for reactive astrogliosis, which is associated with both preclinical and clinical Alzheimer’s disease (AD). Plasma GFAP performed better than cerebrospinal fluid GFAP in discriminating A*β*-positive from A*β*-negative individuals(4). However, a recent study investigating the utility of plasma biomarkers for identifying individuals at the earliest stages for intervention found no association between the levels of NEFL or GFAP in midlife and the later development of dementia(8).

Plasma biomarkers predictive of dementia risk(9) may signify the consequences of the underlying disease processes, but they may not necessarily identify suitable treatment targets. Effective interventions require consideration of causal factors upstream of these protein biomarkers. Causal genetic factors could be expected to influence dementia biomarkers. Established genetic variants associated with large effect-sizes on AD include the two *APOE* missense variants known as *APOE4* and *APOE2*(10, 11) and the two *TREM2* missense variants (R47H and R62H)(12-14), all of which are present with an allele frequency of greater than 0.1% in the European population. Understanding how these AD genetic variants influence specific biomarkers provides insights into gene specific mechanisms. Differential associations of genetic variants with biomarkers inform which cellular processes are preferentially affected by which molecular mechanisms. We therefore investigated how plasma NEFL and GFAP change in relation to these four high-effect AD variants.

## Results

We analyzed plasma proteomics data from 52,618 UKB participants (53.9% female) who did not have a dementia diagnosis at the time of their enrollment, of which 1,605 developed incident all-cause dementia. As expected, logistic regression analyses showed that both NEFL (Beta = 0.78, 95% CI: 0.69 - 0.86, P = 5.4e-70) and GFAP (Beta = 0.89, 95% CI: 0.80 – 0.98, P = 7.2e-85) levels were associated with incident all-cause dementia, adjusting for age, sex, education, and 10 genetic principal components (PCs). While the two biomarkers are positively correlated (r = 0.35, P < 2.2e-16), both remained independently associated with all-cause dementia when they were jointly analyzed in a multiple logistic regression model including all four genetic variants *APOE4, APOE2, TREM2* R47H and R62H (Table 1; Supplementary Table 1).

**Table 1.** Association between plasma biomarkers and genetic risk factors with all-cause dementia in the UK Biobank (n = 52,)

| Variables | Estimate (Beta) | 95% CI | P |
| --- | --- | --- | --- |
| NEFL | 0.658 | 0.552 - 0.762 | 8.9E-35**** |
| GFAP | 0.650 | 0.546 - 0.755 | 3.4E-34**** |
| *APOE4* | 0.878 | 0.787 - 0.969 | 9.3E-80**** |
| *APOE2* | -0.289 | -0.480 - -0.107 | 2.4E-03** |
| *TREM2* R47H | 0.879 | 0.336 - 1.372 | 8.2E-04*** |
| *TREM2* R62H | 0.161 | -0.273 - 0.555 | 4.4E-01 |
| Sex (female) | -0.364 | -0.487 - -0.240 | 7.8E-09**** |
| Age (year) | 0.130 | 0.117 - 0.143 | 1.6E-84**** |
| Education (college) | -0.270 | -0.418 - -0.125 | 3.1E-4*** |

Results from multiple logistic regression model with all-cause dementia as outcome and the independent variables are the plasma proteins as well as the *APOE* and *TREM2* genotypes, adjusting for sex, age at enrollment, education (college vs. no college) and 10 genetic principal components. NEFL: neurofilament light chain; GFAP: glial fibrillary acidic protein. Each protein level was inverse-rank normalized. \*: $P \leq 0.05$ ; \*\*: $P \leq 0.01$ ; \*\*\*: $P \leq 0.001$ ; \*\*\*\*: $P \leq 0.0001$ .

We also investigated whether the two biomarkers in midlife predicted incident dementia, by limiting the analysis to the 28,962 participants who were enrolled at an age <60 years (60-group). Among those, 218 subsequently developed dementia. Given the low allele frequencies of the *TREM2* risk variants and younger age of the participants, only one of the 174 *TREM2* R47H carriers and five out of the 518 R62H carriers developed dementia. Since both risk alleles were reported to increase dementia risk in a large GWAS(14), we aggregated the two variants into one single variable “*TREM2* R47H or R62H”. NEFL and GFAP remained significantly associated with incident dementia in the 60-group (Supplementary Table 2).

Subsequently, we investigated the relationship between the established AD genetic risk variants and the plasma proteins as outcomes in multiple linear regression models including all four genetic variants. We found *APOE4* is significantly associated with greater NEFL (Beta = 0.019, P = 5.3e-5) and greater GFAP levels (Beta = 0.067, P = 3.5e-42) (Table 2). Interestingly, the magnitude of increase of the normalized protein levels with each additional copy of *APOE4* is 3-fold greater for GFAP than for NEFL. When additionally adjusted for GFAP, *APOE4* no longer has any effect on NEFL (Beta=0.005, P=0.27), whereas the effect of *APOE4* on GFAP remains significant when adjusting for NEFL (Beta= 0.063, P=3.8e-39). Contrary to the effect of *APOE4, APOE2* is associated with neither GFAP nor NEFL (Table 2).

**Table 2.** Association between *APOE* and *TREM2* risk variants with plasma proteins.

|  | NEFL |  |  | GFAP |  |  |
| --- | --- | --- | --- | --- | --- | --- |
|  | Estimate (Beta) | 95% CI | P | Estimate (Beta) | 95% CI | P |
| *APOE4* | 0.019 | 0.01 - 0.03 | 5.3E-05**** | 0.067 | 0.06 - 0.08 | 3.5E-42**** |
| *APOE2* | 0.006 | -0.01 - 0.02 | 3.7E-01 | 0.005 | -0.01 - 0.02 | 4.3E-01 |
| *TREM2* R47H | 0.073 | 0.01 - 0.13 | 1.9E-2* | 0.033 | -0.03 - 0.1 | 3.1E-01 |
| *TREM2* R62H | 0.064 | 0.03 - 0.1 | 3.2E-04*** | 0.013 | -0.02 - 0.05 | 4.9E-01 |
| Sex (female) | -0.022 | -0.03 - -0.01 | 8.7E-06**** | 0.154 | 0.14 - 0.16 | 2.6E-198**** |
| Age (year) | 0.035 | 0.03 - 0.04 | <1.0E-300**** | 0.026 | 0.03 - 0.03 | <1.0E-300**** |
| Education (college) | 0.007 | 0 - 0.02 | 1.6E-01 | 0.005 | -0.01 - 0.02 | 3.3E-01 |

Results from two multiple linear regression models with NEFL and GFAP as outcome and the genetic variants as independent variables, adjusted for sex, age, education (college vs no college), and 10 genetic principal components. NEFL: neurofilament light chain; GFAP: glial fibrillary acidic protein. Each protein level was inverse-rank normalized. \*: $P \leq 0.05$ ; \*\*: $P \leq 0.01$ ; \*\*\*: $P \leq 0.001$ ; \*\*\*\*: $P \leq 0.0001$ .

Despite the much lower allele frequencies, both *TREM2* variants R47H and R62H are individually significantly associated with increased NEFL levels (Table 2). Notably, neither of the two variants is associated with GFAP levels. Since both variants showed similar effects on NEFL, they were again aggregated into one variable “*TREM2* R47H or R62H”. The aggregated effect size of *TREM2* variants on NEFL (beta = 0.066, 95% CI: 0.036 – 0.097, P = 2.3e-05) is more than 3-times larger than the effect of *APOE4* on NEFL (Beta = 0.019, 95% CI: 0.010 – 0.028, P = 5.3e-5).

Since dementia biomarkers may be elevated during the prodromal stage of dementia, we performed sensitivity analysis for the association between *APOE4* and the biomarkers stratified by dementia diagnosis. Among participants who developed dementia, *APOE4* is significantly associated with GFAP after controlling for time to dementia diagnosis (Beta = 0.175, P = 2.4e-11) (Supplementary Table 3a, Figure 1a). In this group, we also observed a weak negative correlation between time to dementia and GFAP (Beta = −0.01, P= 4.6e-02) (Supplementary Table 3a). Among participants without incident dementia, *APOE4* remains significantly associated with GFAP (Beta = 0.047, P = 5.5e-21), albeit with a smaller effect (Supplementary Table 3b). Among participants with incident dementia, *APOE4* is not associated with NEFL (Beta = −0.001, P=0.98) after adjusting for time to dementia (Supplementary Table 3c, Figure 1b). Compared to the association with GFAP, time to dementia showed a stronger negative correlation with NEFL (Beta = −0.033, P=1.4e-14) (Supplementary Table 3c). Among participants without incident dementia, *APOE4* showed a very weak correlation with NEFL (Beta = 0.01, P = 3.5e-2) (Supplementary Table 3d, Figure 1b). Among participants without dementia, carrying any of the two *TREM2* variants was associated with higher NEFL levels (Beta = 0.06, P = 5.8e-5) (Supplementary Table 3d), but not among participants who later developed dementia (Supplementary Table 3c).

**Figure 1.**
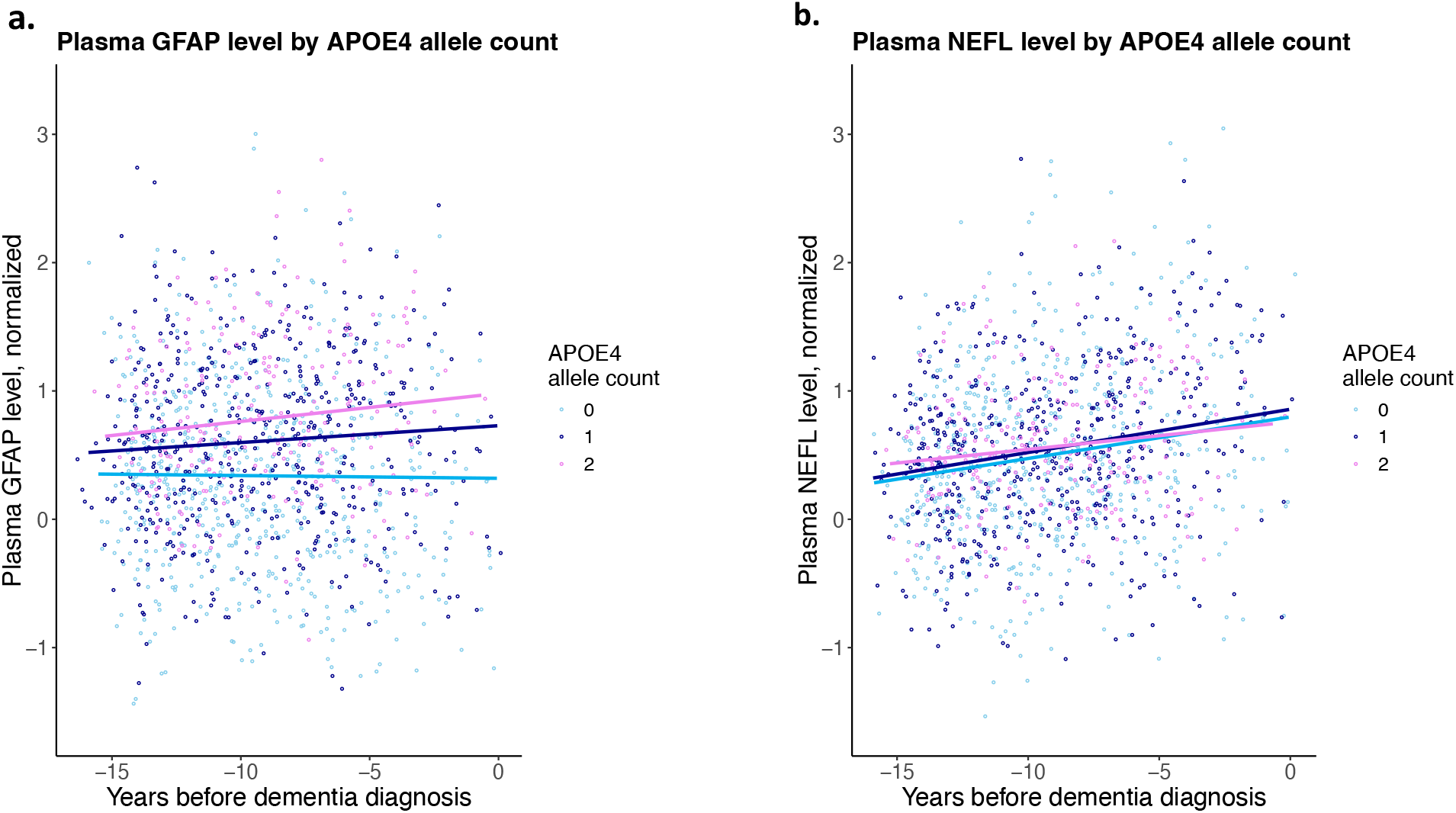
Plasma GFAP and NEFL levels by APOE4 allele counts in participants who developed dementia. **a.** Plasma levels of glial fibrillary acidic protein (GFAP) and **b**. neurofilament light chain (NEFL) stratified by the *APOE4* allele counts in the 1,605 participants who developed incident dementia (633 carried none, 529 carried one, and 212 had two *APOE4* alleles). The x-axis shows the years between enrollment (when the blood sample for the biomarkers were obtained) and the time a dementia diagnosis was entered.

To investigate whether the effects of the genetic variants on the biomarkers depend on age, we performed the analyses stratified by the two age groups: 60- and 60+ (>60 years). While *APOE4* is significantly associated with plasma GFAP in both age groups, the effect size is 4.8-folds greater in the 60+ group compared to the 60-group (Supplementary Table 4). As shown in Figure 2a, carriers of *APOE4* allele showed greater GFAP levels starting in middle age and the effect sizes are greatest in homozygous *APOE4* carriers in both incident dementia and non-dementia groups. When stratified by age, *APOE4* is significantly associated with NEFL only in the 60+ group, not in the 60-group (Supplementary Table 4). We see a similar increase of NEFL with age across *APOE4* genotypes both among incident dementia cases and unaffected participants (Figure 2b). Carriers of either *TREM2* R47H or R62H had greater NEFL levels in both age groups (Supplementary Table 4). *TREM2* risk variants are not associated with GFAP in either age group (Supplementary Table 4, Figure 2c). As shown in Figure 2d, carriers of *TREM2* risk variants without incident dementia have consistently elevated NEFL compared to non-carriers, even in midlife. Interestingly, unlike the relationship between *APOE4* and GFAP, the effect size of TREM2 risk variants does not increase with age (Figure 2d).

**Figure 2.**
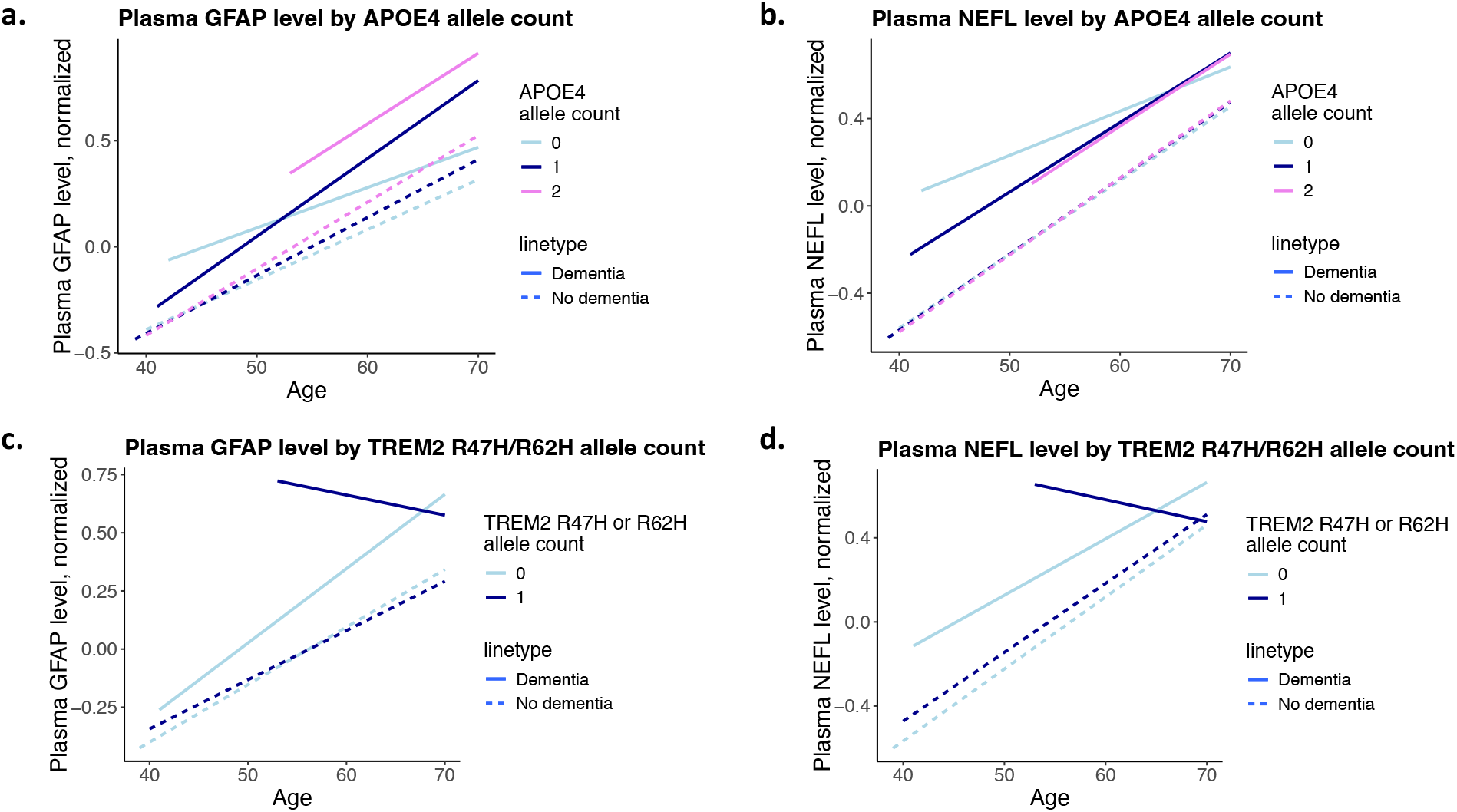
Plasma GFAP and NEFL stratified by APOE4 and TREM2 variants and incident dementia. **a.** plasma glial fibrillary acidic protein (GFAP) and **b**. neurofilament light chain (NEFL) stratified by the APOE4 allele counts; **c**. GFAP and **d**. NEFL levels stratified by *TREM2* R47H or R62H allele counts.

Although not the focus of this study, we observed that being female is associated with greater GFAP levels in both dementia and non-dementia groups and the difference is greater in the dementia group (Beta = 0.27, CI: 0.20 - 0.34, P = 5.4E-14) than in the non-dementia group (Beta = 0.15, CI: 0.14 – 0.16, P = 2.2E-192) (Supplementary Table 3a and 3b). We did not observe a correlation between sex and NEFL in participants with incident dementia, but being female showed a weak correlation with NEFL in those without incident dementia (Supplementary Table 3c and 3d). We performed a sex stratified analysis for the association between APOE4 and GFAP. In both sexes, *APOE4* is associated with greater GFAP (females: beta = 0.073, CI: 0.061 – 0.086, P = 6.0e-30 and males: beta=0.059, CI: 0.045 – 0.073, P=4.9e-17). We did not observe a significant interaction between *APOE4* and sex on GFAP (Beta = 0.016, P = 0.09).

## Discussion

This study investigated the association between high-impact genetic variants in *APOE* and *TREM2* and plasma levels of GFAP and NEFL in the context of incident dementia among the over 50,000 UKB participants. In addition to replicating previous findings that GFAP and NEFL are associated with greater risk for incident dementia, we found that *APOE4* is associated with higher levels of GFAP and NEFL. We also found that *TREM2* variants R47H and R62H are associated with higher NEFL levels, but not GFAP. Furthermore, we found that *APOE4* is more prominently associated with elevated levels of GFAP than NEFL. The *APOE4* effect on GFAP is seen in both middle aged and older participants, and the effect size increases with the number of *APOE4* alleles, regardless of whether the participants developed incident dementia. The effect of *APOE4* on GFAP is also observable in midlife among participants without incident dementia. Contrary to the consistently observed association between *APOE4* and GFAP, no association between *APOE4* and NEFL is observed in participants with incident dementia or in younger participants (<60 years). In addition, while the association between *APOE4* and GFAP remains when NEFL is accounted for, *APOE4* is not associated with NEFL when we adjust for GFAP.

These findings support the hypothesis that *APOE4* exerts its effect over astrocytic activation marked by GFAP elevation that is biologically upstream to neuronal damage represented by NEFL elevation. This interpretation is consistent with astrocytes being the main source of ApoE expression in the brain(15). The selective influence of *APOE4* on GFAP and of *TREM2* variants on NEFL indicates that these genetic risk variants exert their effects on neurodegeneration through different molecular pathways. Astrocytes play a key role in modulating neuroinflammation and regulating brain homeostasis and synaptic transmission(16), and are the main source of ApoE production in the brain(17). Our finding of elevated GFAP in *APOE4* carriers is consistent with experiments in human iPSC-derived astrocytes showing that *APOE4* is a key driver of low-grade chronic inflammation and exacerbated responses to cytokine stimuli that recapitulate the AD environment(18). It is also noteworthy that GFAP has been linked not only to increased brain tau deposition and cognitive decline, but also appears to partially mediate the relationship between brain amyloid burden and tau pathology(19). The lack of association between *TREM2* variants and GFAP is consistent with experimental findings that *TREM2* is expressed in myeloid cells and mainly affects microglial and macrophage function but not astrocytes(20). Interestingly, the protective *APOE2* allele is associated with neither GFAP nor NEFL, suggesting that its protective effect against dementia may follow mechanisms other than astrocytic function. Future research is needed to identify biomarkers influenced by *APOE2*.

Another interesting finding is that unlike the association between *APOE4* and GFAP, for which the magnitude increases with age, the magnitude of association between the *TREM2* variants and NEFL does not increase with age. Since NEFL is a marker of neuronal damage(7), our results suggest that the detrimental effects of *TREM2* risk variants on neurons may present in midlife with a similar effect as in late life. The UKB participants were enrolled between age 40 and 70; future studies that include younger participants are needed to comprehensively investigate the effects of *TREM2* variants across the entire age spectrum. When stratified by incident dementia, *TREM2* risk variants are significantly associated with NEFL only in participants without incident dementia, but not among those who developed dementia. However, owing to the low allele frequency of the *TREM2* variants, only 58 carriers developed incident dementia, limiting our ability to make a conclusive statement. Future studies that include larger numbers of carriers for *TREM2* risk variants are needed to comprehensively interrogate the relationship between *TREM2* variants and NEFL.

The magnitude of association between *APOE4* and GFAP observed in the UKB data aligns with a recent prospective study of over five thousand participants(21). However, our study extends these findings by leveraging a substantially larger sample size (nearly 10-fold greater) and demonstrating the impact of *APOE4* allele dosage, as well as the increasing magnitude of the *APOE4* effect with advancing age. Furthermore, while GFAP levels rise with age across the entire cohort, our results indicate that the excessive GFAP elevation observed in *APOE4* carriers begins before the age of 60.

We observed significant associations of both GFAP and NEFL with future all-cause dementia in the UKB participants. Our finding is consistent with previous reports that plasma GFAP was higher in cognitively normal older adults with high brain amyloid-β load, indicating that GFAP is associated with preclinical AD(4, 22). We show that both biomarkers remained significantly associated with future dementia when we restricted the analysis to participants younger than 60 years at the time of enrollment. A previous study reported an association between plasma NEFL and GFAP levels and incident dementia in late-life, but not midlife(8). Our findings replicate the late-life association but, in contrast, demonstrate that midlife levels of these biomarkers are associated with incident dementia. This suggests that preventative interventions for individuals carrying major genetic risk factors may need to be initiated during midlife.

Finally, we noticed that being female is associated with greater GFAP levels and *APOE4* is associated with elevated GFAP in both sexes. Interestingly, a previous report found that females have higher blood GFAP levels than males after traumatic brain injuries and that higher GFAP was associated with functional limitations only in females(23). We did not observe a significant interaction between *APOE4* and sex on GFAP. However, larger sample sizes might be required to identify such interactions.

This study has several limitations. First, the UKB blood samples were collected at a single time point at enrollment, precluding longitudinal tracking of biomarker changes within individuals. Nevertheless, the broad age range of participants (40-70 years) at enrollment allows for investigation of age-related biomarker changes across midlife to later life, as demonstrated here. Future studies incorporating younger individuals are necessary to determine whether these genetic effects on NEFL and GFAP are present from a young age or emerge as an early feature of prodromal dementia. Second, the lack of plasma amyloid beta and phosphorylated tau data in the UKB proteomics dataset currently prevents investigation of the relationship between genetic factors and these established AD markers. Third, after controlling for NEFL and GFAP, we found no significant associations between *APOE4, APOE2* and *TREM2* variants and incident dementia in participants younger than 60. This is likely attributable to the lower incidence of dementia in this younger age group, as the oldest participants at the time of data extraction were only in their mid-70s. Fourth, due to the sample size limitation, we did not investigate rarer variants in *APOE, TREM2*, and in other AD risk genes. Future proteomics data of all 500,000 UKB participants could enable investigations of very rare high-effect variants. Finally, future investigations could include additional risk factors such as hypertension, diabetes, cardiovascular disorders, and environmental factors.

In conclusion, key dementia blood biomarkers GFAP and NEFL are differentially impacted by genetic risks for AD, pointing to gene specific pathways to neurodegeneration. *APOE4* influences GFAP levels (astrocyte activation) in an age- and dosage-dependent manner. In contrast, *TREM2* variants affect NEFL (neuronal damage) without discernable impact on GFAP. Both GFAP and NEFL in midlife predict future dementia risk, indicating interventions may need to start in midlife. More research is needed to link each individual dementia genetic risk factor with downstream biomarker changes to fully understand how genetic risks lead to dementia.

## Methods

### The UK Biobank data

The UK Biobank (UKB) is a prospective population-based cohort study of ~500,000 participants from the United Kingdom, enrolled at an age between 37 and 69 years between 2006–2010(24). Genome-wide genotyping, levels of blood biomarkers, and diagnose codes through linkage to electronic health records (EHR) are available. All participants provided informed consent for approved scientific use of their UKB data (https://biobank.ndph.ox.ac.uk/showcase/). The present study is approved under application number 84323.

Details of the proteomics data from the UKB have been extensively described before (5, 6, 25). Proteomic profiling was done on plasma samples from 54,219 UKB participants using the anti-body-based Olink Explore 3072 Proximity Extension Assay as part of the UK Biobank Pharma Proteomics Project (UKB-PPP) capturing 2,923 unique proteins. Documentations of the UKB-PPP can be found at https://biobank.ndph.ox.ac.uk/showcase/label.cgi?id=1839. We obtained plasma NEFL and GFAP levels as normalized proteomic measures from the UKB-PPP project that have been processed through a quality-control process(5) (see also: https://bi-obank.ndph.ox.ac.uk/ukb/ukb/docs/Olink_1536_B0_to_B7_Normalization.pdf).

The genotypes for the established AD risk variants *APOE4* (rs429358), *APOE2* (rs7412) and *TREM2* R47H (rs75932628) and R62H (rs143332484) are obtained from whole-exome sequencing data(26). Variant calling and quality control steps have been described previously(26). We used the genomic tools that are implemented on the UK Biobank Research Analysis Platform (UKB-RAP) including bgenix(27) and PLINK 2.0(28) to extract individual genotypes of the variants of interest by their genomic positions.

The diagnosis information for dementia and time of diagnosis entries are obtained from the data category 1712 “First Occurrences” (UKB data accessed on April 2^nd^ 2024). The diagnosis data are based on international classification of diseases 10th revision (ICD-10) and were derived by cross-linking health registers, including primary care, hospital inpatient, and death registers. All-cause dementia cases were aggregated from AD, vascular dementia, dementia classified elsewhere, and unspecified dementia defined as ICD-10 codes F00, F01, F02, F03, F04, G30, G31, and G32, as we described before(29).

### Statistical analysis

We first evaluated data integrity by investigating the association between NEFL and GFAP with dementia as the dependent variable separately in a logistic regression model adjusting for sex, age at recruitment, education (college or above vs no college) and 10 genetic principal components. Next, we included both NEFL and GFAP and the genetic variants *APOE4, APOE2, TREM2* R47H and R62H as additional covariates to assess the effects of NEFL and GFAP on dementia that are independent from each other and from these genetic variants. Sensitivity analyses were performed using analysis stratified by age group, i.e. <60 (60-) and >=60 years (60+) at enrollment.

To investigate the relationship between the four genetic variants and the biomarkers, we used multiple linear regression using each biomarker as the dependent variable and the genetic risk factors as independent variables, adjusting for sex, age at recruitment, education and 10 genetic principal components. We also performed sensitivity analysis by stratifying the participants into two groups – those with and without incident dementia.

Since the goal of this study is to investigate the relationship between established AD genetic variants APOE4, APOE2, TREM2 R47H and R62H with established AD biomarkers NEFL and GFAP, significance is defined as P<0.05. All statistical analyses were performed using R version 4.4.1(30).

## Data Availability

UK Biobank data are available through thelink below

https://www.ukbiobank.ac.uk/enable-your-research/apply-for-access

## Author Contributions

Conceptualization: YFH, CD

Data curation: YFH

Formal analysis: YFH

Funding acquisition: YFH

Methodology: YFH, WL, AG

Project administration: YFH

Resources: YFH, JK

Supervision: WL, YM, JK, LG

Validation: WL

Writing – original draft: YFH

Writing – review & editing: all authors

All authors read and approved the final version of the manuscript.

## Data Sharing Statement

UK Biobank data are available through application at: https://www.ukbiobank.ac.uk/enable-your-research/apply-for-access

## Acknowledgments

YFH received founding from NIH/NIA (K08AG054727).

