## Supplemental tables for "*APOE* and *TREM2* variants differentially influence glial fibrillary acidic protein and neurofilament light in plasma of UK Biobank participants"

**Supplementary Table 1. Number of participants with dementia genetic risk variants in the UK Biobank proteomics data**

| | Numer of alleles | Non-dementia | Dementia |
|---|---|---|---|
| *APOE4* | 0 | 30,927 | 633 |
| | 1 | 11,101 | 529 |
| | 2 | 1,078 | 212 |
| | Missing | 7,907 | 231 |
| *APOE2* | 0 | 38,214 | 1,304 |
| | 1 | 6,756 | 124 |
| | 2 | 349 | 10 |
| | Missing | 5,694 | 167 |
| *TREM2* R47H | 0 | 48,548 | 1,500 |
| | 1 | 298 | 26 |
| | 2 | 0 | 2 |
| | Missing | 2,167 | 77 |
| *TREM2* R62H | 0 | 47,927 | 1,498 |
| | 1 | 915 | 29 |
| | 2 | 4 | 1 |
| | Missing | 2,167 | 77 |

Number of alleles: number of minor alleles of the variants carried by a participant. Non-dementia: Participants who did not have an incident dementia diagnosis by 4/2/2024. Dementia: Participants did not have a dementia diagnosis at enrollment, but had a dementia diagnosis by 4/2/2024.

**Supplementary Table 2. Association between plasma biomarkers and genetic risk factors with all-cause dementia among participants <60 years old at enrollment (n= 28,962)**

| | Estimate | 95% CI | P |
|---|---|---|---|
| NEFL | 0.983 | 0.76 - 1.2 | 5.9E-19**** |
| GFAP | 0.669 | 0.4 - 0.92 | 4.3E-07**** |
| *APOE4* | 0.259 | -0.04 - 0.54 | 7.7E-02 |
| *APOE2* | -0.382 | -0.92 - 0.09 | 1.3E-01 |
| *TREM2* R47H or R62H | 0.045 | -1.01 - 0.86 | 9.2E-01 |
| Sex (female) | -0.556 | -0.88 - -0.23 | 7.7E-04*** |
| Age (year) | 0.065 | 0.03 - 0.1 | 1.8E-04*** |
| Education (college) | -0.384 | -0.75 - -0.04 | 3.3E-02* |

Results from the multiple logistic regression model among participants enrolled at age <60 years, with all-cause dementia as depedent variable and the independent varianbles are the plasma proteins as well as the *APOE* and *TREM2* genotypes. Since only one out of 174 R47H carriers and five out of 518 R62H carriers developed dementia, the two *TREM2* variants are aggregated into one single variable "*TREM2* R47H or R62H". The model adjusted for sex, age at enrollment, education (college vs. no college) and 10 genetic principal components. NEFL: neurofilament light chain; GFAP: glial fibrillary acidic protein. Each protein level was inverse-rank normalized. PC: genetic principal components. *: P<=0.05; **: P<=0.01; ***: P<=0.001; ****: P<=0.0001.

**Supplementary Table 3a. Association of genetic variants with GFAP in participants with incident dementia, adjusting for time to dementia (n=1,605)**

| | Beta | 95% CI | P |
|---|---|---|---|
| Time to dementia (year) | -0.010 | -0.020 - 0 | 4.6E-02* |
| *APOE4* | 0.175 | 0.124 - 0.226 | 2.4E-11**** |
| *APOE2* | 0.026 | -0.084 - 0.135 | 6.5E-01 |
| *TREM2* R47H or R62H | -0.005 | -0.177 - 0.167 | 9.5E-01 |
| Sex (female) | 0.274 | 0.203 - 0.344 | 5.4E-14**** |
| Age at recruitment | 0.026 | 0.019 - 0.033 | 1.1E-12**** |
| Education | 0.048 | -0.037 - 0.133 | 2.7E-01 |

**Supplementary Table 3b. Association of genetic variants with GFAP in participants without incident dementia (n=51,013)**

| | Beta | 95% CI | P |
|---|---|---|---|
| *APOE4* | 0.047 | 0.037 - 0.057 | 5.5E-21**** |
| *APOE2* | 0.007 | -0.006 - 0.020 | 3.1E-01 |
| *TREM2* R47H or R62H | 0.012 | -0.020 - 0.044 | 4.6E-01 |
| Sex (female) | 0.153 | 0.142 - 0.163 | 2.2E-192**** |
| Age at recruitment | 0.025 | 0.025 - 0.026 | <1.0E-300**** |
| Education | 0.006 | -0.004 - 0.017 | 2.4E-01 |

**Supplementary Table 3c. Association of genetic variants with NEFL in participants with incident dementia, adjusting for time to dementia (n=1,605)**

| | Estimate | | P |
|---|---|---|---|
| Time to dementia (year) | -0.035 | -0.043 - -0.026 | 5.5E-15**** |
| *APOE4* | -0.001 | -0.045 - 0.044 | 9.8E-01 |
| *APOE2* | 0.059 | -0.037 - 0.156 | 2.3E-01 |
| *TREM2* R47H or R62H | 0.026 | -0.126 - 0.178 | 7.4E-01 |
| Sex (female) | 0.030 | -0.032 - 0.092 | 3.4E-01 |
| Age at Recruitment | 0.026 | 0.02 - 0.032 | 3.3E-17**** |
| Education | 0.017 | -0.058 - 0.092 | 6.6E-01 |

**Supplementary Table 3d. Association of genetic variants with NEFL in participants without incident dementia (n=51,013)**

| | Beta | 95% CI | P |
|---|---|---|---|
| *APOE4* | 0.010 | 0.001 - 0.02 | 3.5E-02* |
| *APOE2* | 0.006 | -0.007 - 0.018 | 3.6E-01 |
| *TREM2* R47H or R62H | 0.064 | 0.033 - 0.095 | 5.8E-05**** |
| Sex (female) | -0.021 | -0.03 - -0.011 | 2.9E-05**** |
| Age at Recruitment | 0.034 | 0.033 - 0.035 | <1.0E-300**** |
| Education | 0.009 | -0.001 - 0.02 | 7.8E-02 |

Results from sensitivity analyses stratifying by whether the participants developed incident dementia. For all multiple regressions, the dependent variables was GFAP or NEFL and independent variables are the genetic variants, adjusted for sex, age and education (college vs no college) and 10 genetic principal components. In the dementia sub-group, the regression model also included time-to-dementia defined as years between the enrollment and the demenia. GFAP: glial fibrillary acidic protein. PC: genetic princical components. Each protein level was inverse-rank normalized. Regression results for GFAP (**a.**) and NEFL (**c.**) in participants who developed incident all-cause dementia. Regression results for GFAP (**b.**) and NEFL (**d.**) in participants without incident all-cause dementia. *: P<=0.05; **: P<=0.01; ***: P<=0.001; ****: P<=0.0001.

**Supplementary Table 4. Association of genetic risk factors with plasma biomarkers stratified by age at enrollment of participants**

| | GFAP, 60- (n=28,962) | | | GFAP, 60+ (n=23,656) | | | NEFL, 60- (n=28,962) | | | NEFL, 60+ (n=23,656) | | |
|---|---|---|---|---|---|---|---|---|---|---|---|---|
| | Estimate | 95% CI | P | Estimate | 95% CI | P | Estimate | 95% CI | P | Estimate | 95% CI | P |
| *APOE4* | 0.023 | 0.01 - 0.035 | 4.3E-04**** | 0.111 | 0.096 - 0.125 | 1.5E-49**** | 0.007 | -0.006 - 0.019 | 2.9E-01 | 0.030 | 0.016 - 0.044 | 1.8E-05**** |
| *APOE2* | -0.002 | -0.018 - 0.015 | 8.3E-01 | 0.010 | -0.01 - 0.03 | 3.4E-01 | 0.010 | -0.007 - 0.026 | 2.6E-01 | -0.001 | -0.02 - 0.018 | 9.2E-01 |
| *TREM2* R47H or R62H | 0.037 | -0.004 - 0.077 | 7.5E-02 | -0.007 | -0.057 - 0.043 | 7.8E-01 | 0.078 | 0.037 - 0.118 | 1.7E-04*** | 0.052 | 0.005 - 0.098 | 2.9E-02* |
| Sex (female) | 0.122 | 0.109 - 0.134 | 7.1E-77**** | 0.199 | 0.183 - 0.215 | 1.5E-132**** | -0.028 | -0.04 - -0.015 | 1.9E-05**** | -0.011 | -0.026 - 0.003 | 1.3E-01 |
| Age (year) | 0.021 | 0.02 - 0.022 | 7.8E-289**** | 0.035 | 0.032 - 0.038 | 7.9E-139**** | 0.031 | 0.03 - 0.032 | <1.0E-300**** | 0.042 | 0.04 - 0.045 | 4.1E-226**** |
| Education (college) | 0.014 | 0.001 - 0.027 | 4.1E-02* | 0.001 | -0.017 - 0.019 | 9.4E-01 | 0.020 | 0.006 - 0.033 | 3.4E-03** | -0.007 | -0.023 - 0.01 | 4.5E-01 |

Multiple regression analysis stratified by age groups into <60 years (60-) and >=60 years (60+) at enrollment. For all regression models, the outcome was GFAP or NEFL and explanatory variables are the genetic variants, adjusted for sex, age and education (college vs no college) and 10 genetic principal components. NEFL: neurofilament light chain; GFAP: glial fibrillary acidic protein. Each protein level was inverse-rank normalized. *: P<=0.05; **: P<=0.01; ***: P<=0.001; ****: P<=0.0001.
